# Temporal genomic analysis of *Plasmodium falciparum* reveals increased prevalence of mutations associated with delayed clearance following treatment with artemisinin-lumefantrine in Choma District, Southern Province, Zambia

**DOI:** 10.1101/2024.06.05.24308497

**Authors:** Abebe A. Fola, Tamaki Kobayashi, Timothy Shields, Harry Hamapumbu, Michael Musonda, Ben Katowa, Japhet Matoba, Jennifer C. Stevenson, Douglas E. Norris, Philip E. Thuma, Amy Wesolowski, William J. Moss, Jonathan J. Juliano, Jeffrey A. Bailey, Southern and Central Africa International Center of Excellence for Malaria Research (ICEMR)

**Author notes:** Corresponding Author Abebe A. Fola, 55 Claverick Street., RM#314A, Department of Pathology and Laboratory Medicine, Brown University. Co-senior author (Jonathan J. Juliano, Jeffrey A. Bailey).

## Abstract

The emergence of antimalarial drug resistance is an impediment to malaria control and elimination in Africa. Analysis of temporal trends in molecular markers of resistance is critical to inform policy makers and guide malaria treatment guidelines. In a low and seasonal transmission region of southern Zambia, we successfully genotyped 85.5% (389/455) of *Plasmodium falciparum* samples collected between 2013-2018 from 8 spatially clustered health centres using molecular inversion probes (MIPs) targeting key drug resistance genes. Aside from one sample carrying K13 R622**I**, none of the isolates carried other World Health Organization-validated or candidate artemisinin partial resistance (ART-R) mutations in K13. However, 13% (CI, 9.6-17.2) of isolates had the AP2MU S160**N** mutation, which has been associated with delayed clearance following artemisinin combination therapy in Africa. This mutation increased in prevalence between 2015-2018 and bears a genomic signature of selection. During this time period, there was an increase in the MDR1 N**F**D haplotype that is associated with reduced susceptibility to lumefantrine. Sulfadoxine-pyrimethamine polymorphisms were near fixation. While validated ART-R mutations are rare, a mutation associated with slow parasite clearance in Africa appears to be under selection in southern Zambia.

## Introduction

*Plasmodium falciparum* malaria remains an overwhelming problem in Africa, where approximately 90% of global cases and deaths occur, affecting primarily children younger than five years of age and pregnant women.^1^ Effective antimalarial drugs are a cornerstone for both malaria treatment and prevention. The emergence, continued evolution, and spread of antimalarial drug resistance will undermine ongoing control and elimination efforts.^2–4^ Of particular concern is evolving artemisinin and artemisinin combination therapy (ACT) resistance in eastern Africa.^5–7^ Resistance phenotyping of parasites *ex vivo* and clinical therapeutic efficacy studies (TESs) are useful tools for assessing resistance,^8^ however, phenotypic characterization is technically challenging, costly, labor-intensive, and not as scalable as widespread genomic surveillance. Importantly, parasite clearance curves in TES are less informative in African regions where most of infections consist of multiple parasite strains,^9,10^ meaning that resistant parasites are likely to share their host with sensitive parasites, obscuring detection, particularly early during the spread of resistance.^10,11^ Molecular surveillance of *P. falciparum* parasites is a powerful tool for early detection of emergence and monitoring the spread of antimalarial drug resistance for known resistance mutations and genes.

Currently, the World Health Organization (WHO) recommends artemisinin combination therapy (ACT) as first-line treatment in all African countries.^4^ Sulfadoxine-pyrimethamine (SP) is the recommended first line drug for the intermittent preventive treatment for pregnant women (IPTp) and infants (IPTi) living in high-transmission areas of Africa. However, studies provide evidence that *P. falciparum* has developed resistance to most available antimalarial drugs including SP and ACTs.^3,12–15^ For ACTs, TES results from several African countries have shown early signs of treatment failure of the ACT partner drugs by day 28 or 42.^16,17^ Moreover, recent studies showed increased frequency of WHO-validated/candidate mutations (R561**H**, A675**V** and C469**Y**) in the *pfkelch13* (K13) gene in Rwanda, Tanzania and Uganda ^6,18,19^ and 622**I** in Eritrea and Ethiopia.^5,7^ The emergence of artemisinin partial resistance (ART-R) in Africa, likely heralds the future emergence of partner drug resistance and the eventual clinical failure of ACTs, as seen in SE Asia.^20,21^

In Zambia, despite intensified control interventions, *P. falciparum* malaria remains endemic with high heterogeneity in transmission at the provincial level.^22^ Chloroquine was the frontline drug along with SP until 2002 when ACTs were adopted, with AL the predominant first line treatment nationwide. SP is still employed for IPTp.^23^ The emergence and spread of antimalarial drug resistance in neighbouring countries, particularly artemisinin partial resistance,^18,24^ is an additional threat to national malaria control and elimination. In areas of low transmission, premunition will decrease ^25^ and may enhance the emergence of drug resistance, as a greater proportion of infected individuals seek treatment thereby exposing the parasite population to greater drug pressure.^26^ And once evolved, drug resistant strains that can survive treatment can potentiate malaria resurgence and epidemics.^27,28^ Strong molecular malaria surveillance using multiplexed molecular genotyping and data analysis tools is essential to track the early emergence and spread of antimalarial drug resistance mutations to mitigate its impact.

Despite these concerns, temporally informative surveillance of antimalarial drug resistance in Zambia is limited. While validated mutations associated with ART-R in the K13 gene have not been detected, parasites carrying mutations that confer SP resistance are highly prevalent in the country.^23,29^ Therefore, serial or continuous collections of samples can help us better understand trends in antimalarial drug resistance. Here, we successfully genotyped 389 *P. falciparum* samples collected from 2012-2018 from 8 health centres in Choma District, a low malaria transmission zone in, Southern Province, Zambia, using 815 molecular inversion probes (MIPs) targeting 14 key *P. falciparum* drug resistance genes to determine temporal trends in resistance mutations. In addition, we performed whole genome sequencing (WGS) of 28 samples collected in 2019 from the same area to assess genomic signatures of selection at key antimalarial resistance polymorphisms. Together, the findings suggest that *P. falciparum* parasites in Zambia are under strong selective pressure with an increase in prevalence and genomic evidence of positive selection at loci that may have an impact on ACT effectiveness.

## Results

### K13 gene polymorphism

From the 389 (85%) samples successfully sequenced, we first assessed the prevalence of WHO validated or candidate mutations associated with ART-R. One sample from Mangunza Health Centre in 2016 carried the validated ART-R K13 622**I** pure mutant. This mutation has become common in the Horn of Africa and was recently validated in TES in Eritrea.^5,7,30^ Five novel mutations, C532**S**, A578**S**, Q613**E**, D680**N** and G718**S**, - within the Kelch 13 propeller domain and 6 non-synonymous (NS) mutations outside the Kelch 13 propeller domain were found at low frequencies (**Fig S1A**). The A578**S** mutation is close to the validated mutation C580**Y**, which is associated with ART-R in south-east Asia. We also identified the common K189**T** polymorphism in Africa that is not associated with resistance in 13.2% (95% CI, 9.5-17.4) of samples. The Kelch 13 mutations R561**H**, A675**V**, and C469**Y** recently reported in Rwanda, Tanzania, and Uganda^31^ were not identified. None of the samples carried mutations that were associated with the genetic backbone for ART-R in south-east Asia such as apicoplast ribosomal protein s10 (ARPS10 V127**M**), ferredoxin (FD D193**Y**), *P. falciparum* multidrug resistance 2 transporter (MDR2 T484**I**), putative phosphoinositide-binding protein (PIB7 C1484**F**), *P. falciparum* protein phosphatase (PPH V1157**L**), and *P. falciparum* chloroquine resistance transporter (CRT N326**S** and CRT I356**T**). The prevalence of other key mutations identified in this study can be found in **Fig S1B** and **Table S1**.

### Emerging S160N mutation in *Plasmodium falciparum* protein complex 2 mu subunit (AP2MU) gene

We assessed polymorphisms in the AP2MU gene and found an increasing prevalence of 160**N**, a mutation putatively associated with ACT delayed clearance in Africa^32,33^, from 0% in 2012 and 2013 to 16.3% in 2018 (**Fig 1A**). The 160**N** mutation was detected in samples from all health facilities with spatial heterogeneity (range 6.6 % in Kabanze to 17.6 % in Kamwanu) (**Fig 1B**). All parasites carrying the 160**N** mutation also carried the CRT wild-type (C72, V73, M74, N75, K76) haplotype suggesting reversal to chloroquine sensitive strains in Zambia.

**Figure 1.**
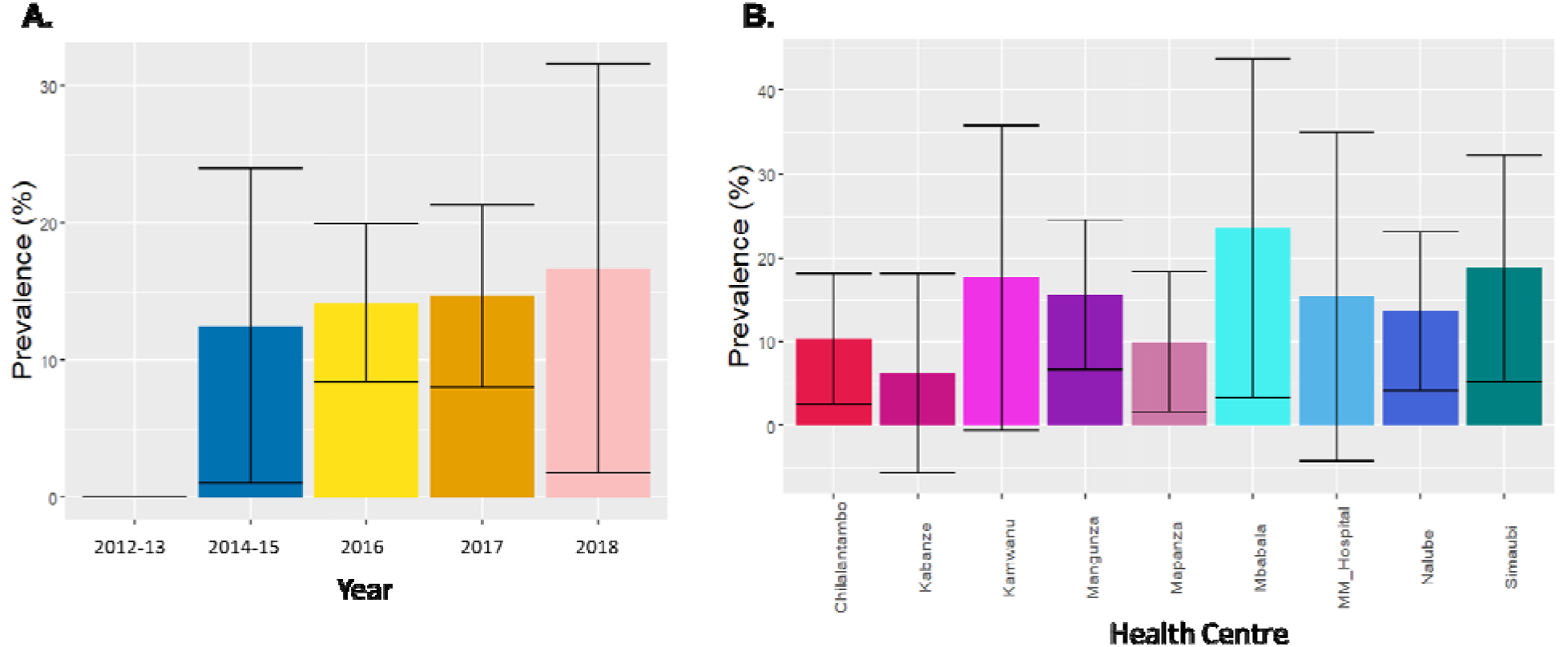
Spatio-temporal trends of AP2MU S160N mutation. **A)** Temporal prevalence of S160**N** mutation in Choma District, Southern Province, Zambia. **B)** Prevalence of the S160**N** mutation at the health facility level. Barplot showing the mean values with standard deviation as error.

### *P. falciparum* ATPase 6 (ATP6) and *P. falciparum* ubiquitin-specific protease 1 (UBP1) gene polymorphisms

Of the four mutations L263**E**, E431**K**, A623**E**, S769**N** in the ATPase 6 gene previously associated with increased artemether IC_50_, 13.1% (CI, 9.4-16.5) of genotyped samples had the E431**K** mutation. One additional mutation, N569**E**, was found at a high frequency (31.1%, CI, 26.5-37.2) with less temporal variation. We detected additional nonsynonymous (NS) mutations in the ATPase 6 gene at low frequency (**Fig 2A**). In UBP1, two key mutations D1525**E**, and E1528**D**, also are associated with delayed clearance of parasites in Africa.^33,34^ These mutations were found at 4.7% (CI, 2.4-7.4) and 8.7% (CI, 5.4-11.9) prevalence, respectively. Genotyped samples also had an additional 28 different NS mutations in UBP1 gene with variable prevalence (ranges 2 to 100 %) (**Fig 2B**).

**Figure 2.**
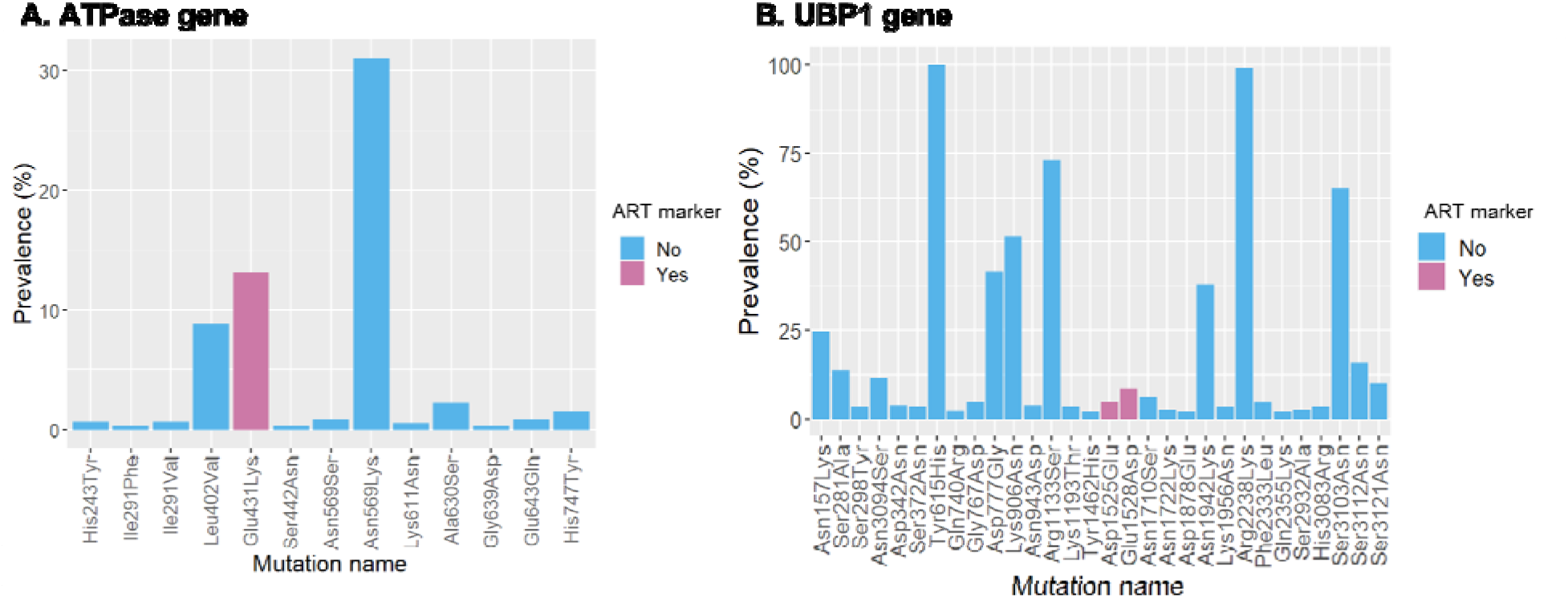
Non-synonymous polymorphism in ATPase 6 (A) and UBP1 (B) genes. The bar plots show the prevalence and colours whether the mutation is the artemisinin resistant marker or not within each gene.

### Increased prevalence of multi drug resistance 1 (MDR1) NFD haplotype

Mutations in the *P. falciparum* multi-drug resistance gene 1 (MDR1), particularly MDR1 N86, 184**F**, and D1246 (N**F**D haplotype), are associated with decreased sensitivity to lumefantrine.^35,36^ Overall, 41% (CI, 34.4-45.1) of genotyped samples carried the Y184**F** mutant allele (**Fig 3A, Table S1**) with some spatial variation (**Fig S2**). Overall, 38.5% (121/314) of samples carried the N**F**D haplotype with increasing prevalence over time (**Fig 3B**). Only 3 genotyped samples carried the N86**Y** mutation that has been associated with enhanced resistance to chloroquine, further supporting reversal to chloroquine-sensitive parasites in southern Zambia.

**Figure 3.**
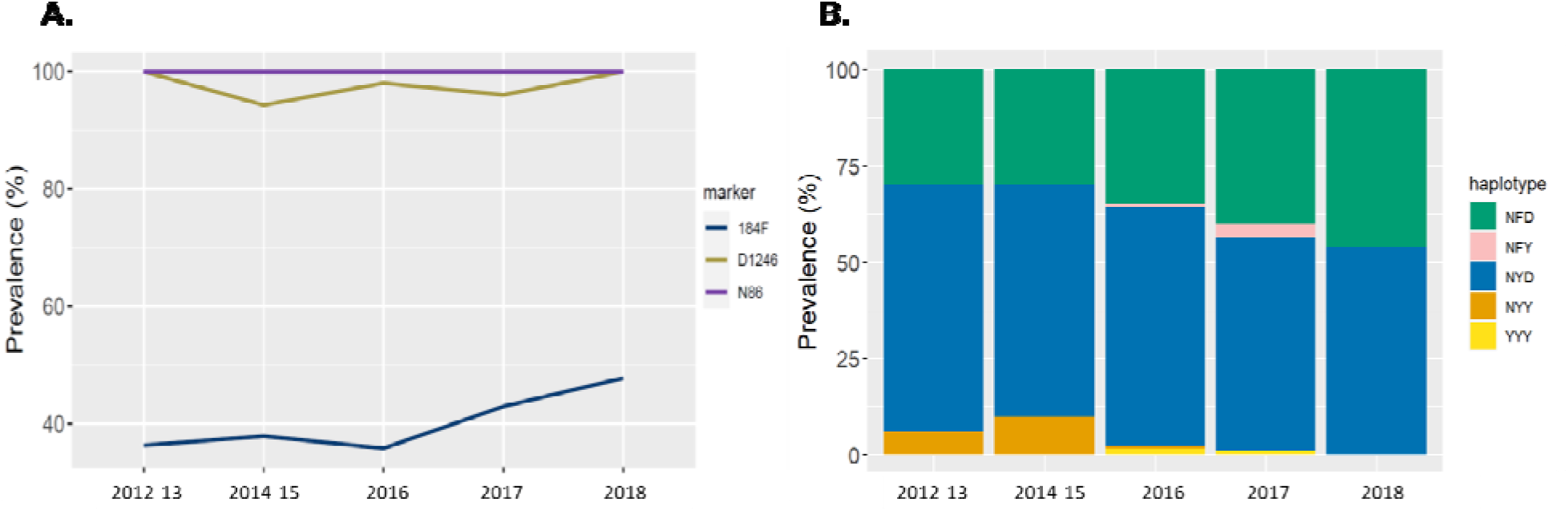
Temporal trends of MDR1 N86, 184F and D1246 mutations in Choma District, Southern Province, Zambia. Colors indicate genotypes (mutant or wildtype) (A) and haplotypes combination of N86/184F/D1246 at MDR1 gene (B).

### Spatio-temporal trend of DHFR and DHPS mutations

Resistance to sulfadoxine-pyrimethamine (SP) in *P. falciparum* is determined by multiple mutations in two genes encoding dihydrofolate reductase (DHFR) related to pyrimethamine and dihydropteroate synthase (DHPS) related to sulfadoxine (**Supplementary Table 1)**. Overall, 91.8 % (290/316) of genotyped samples carried point mutations at codons DHFR 51**I**, 59**R**, and 108**N** (**IRN**-triple mutant). Also, 80.8 % (237/293) of genotyped samples carried point mutations at codons DHPS 437**G** and 540**E** (**GE**-double mutant). Overall, 78.8% (220/279) of samples carried five mutations at DHFR 51I/59R/108N and DHPS 437G/540E (**IRNGE**), the quintuple mutant (**Fig 4A**), with an increase in prevalence in recent years (**Fig 4B**). Only two sequenced samples (one from Nalube Rural Health Center in 2016 and the other is from Mbabala Rural Health Center in 2017) carried the DHPS 581**G** mutation, which increases SP resistance in combination with the DHFR-DHPS quintuple mutant haplotype. None of the isolates had the DHFR 164**L** mutation that confers high-level resistance to SP.

**Figure 4.**
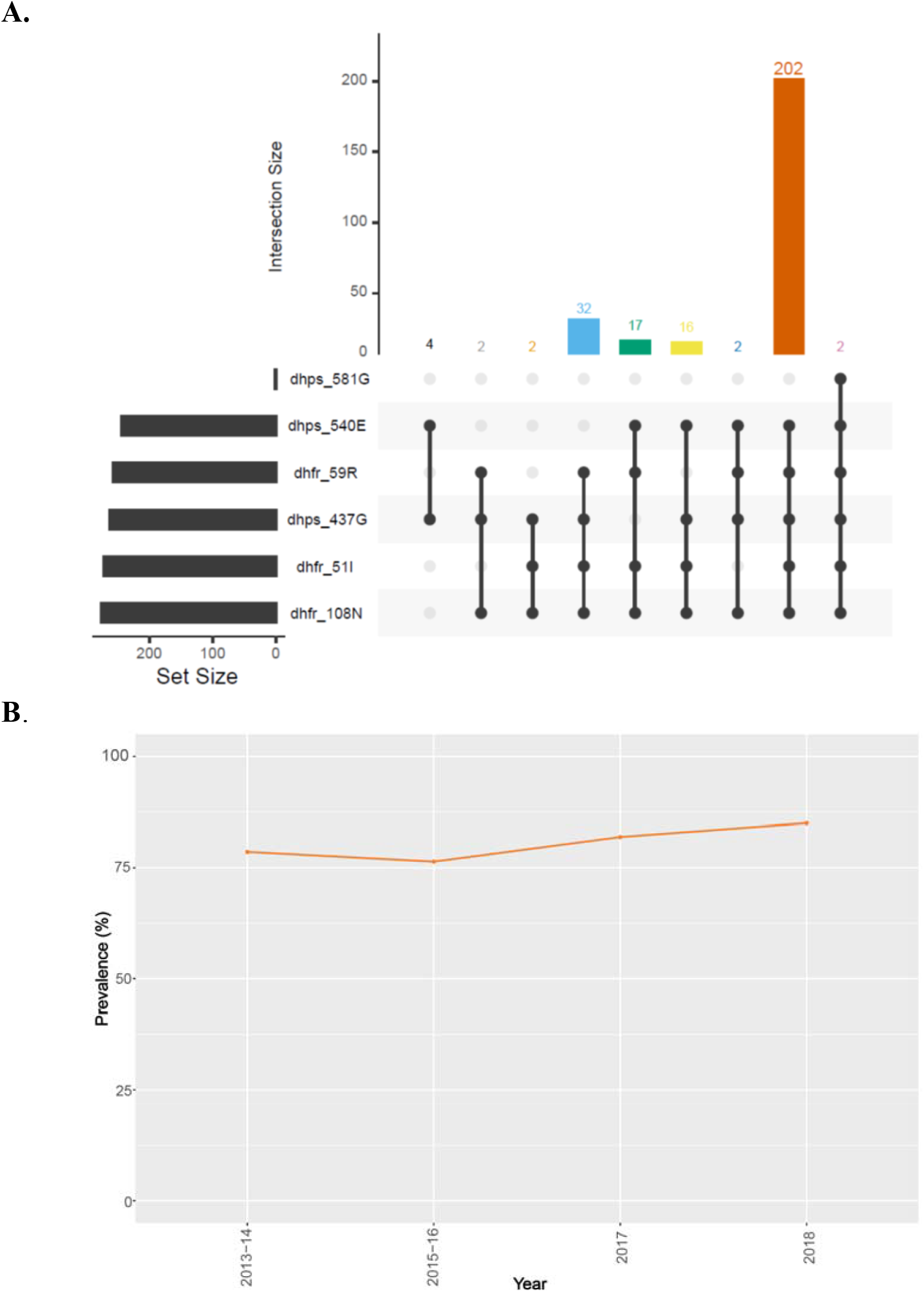
Prevalence of DHFR/DHPS genotypes in Choma District, southern Zambia. **A**) UpSet plots showing the number of times each combination of mutations was seen for Pfdhfr and Pfdhps genes IRNGE quintuple mutant parasites are the most common. B) Temporal trends of IRNGE quintuple mutant across Choma District, Southern Province, Zambia. IRNGE = DHFR 51I/59R/108N and DHPS 437G/540E genotypes.

### Spatio-temporal trend of S258L mutation in putative *P. falciparum* amino acid transporter (AAT1) gene

The 258**L** mutation in the AAT1 gene was found at high frequency (range 89.4-100%) across different health facilities and increased in frequency at end of study period (**Fig 5**). However, none of the isolates carried AAT1 313**S**, a key mutation that augments CQ resistance when it occurs together with 258**L**.^37^ Importantly, all isolates carried CRT K76 wild-type across the years suggesting CQ sensitivity was maintained in Zambia.

**Figure 5.**
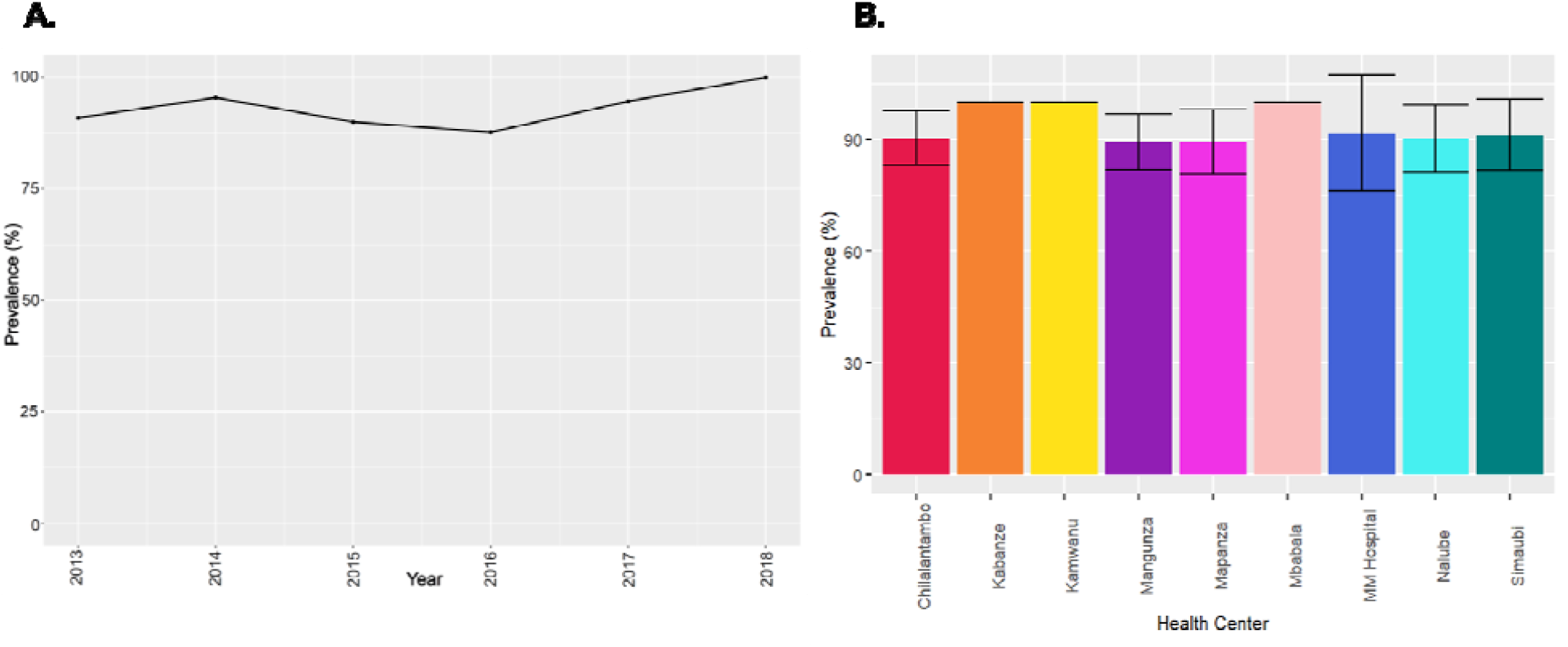
Spatio-temporal trends of AAT1 S258L mutation Choma District, Southern Province, Zambia. **A)** Temporal prevalence S258**L** mutation in Choma District, Southern Province, Zambia. **B)** Prevalence of the S258**L** mutation at the health facility level. Bar plot showing the mean values with standard deviation as error.

### Haplotype-based signatures of selection in southern Zambia

Using WGS and iHS statistics analysis, we identified several loci under positive selection in the parasite population (**Fig 6A**). Except for the AP2MU gene, no selection signals were observed around the five known drug resistance genes that include CRT, MDR1, DHFR, DHPS, and K13. The EHH analysis detected significantly elevated haplotype homozygosity associated with the mutant allele AP2MU 160**N (Fig 6B and C**, red line), which may be driven by ACT pressure resulting in a selective sweep. Moreover, the iHS analysis identified the selection signals for important genes such *PfEMP1* (*P. falciparum* erythrocyte membrane protein ) that has been shown to be involved in sequestration and antigenic variation, other genes implicated in roles for immune evasion, aggregation, or cytoadherence to microvasculature repetitive interspersed families of polypeptides (RIFINs), subtelomeric variable open reading frame (STEVOR), vaccine candidate *P. falciparum* apical membrane antigen 1 (AMA1) and other genes (see list of genes under selection in **Tables S2**). Selection across these loci may be indicative of decreasing effective population size and increasing clonal spread in the population.

**Figure 6.**
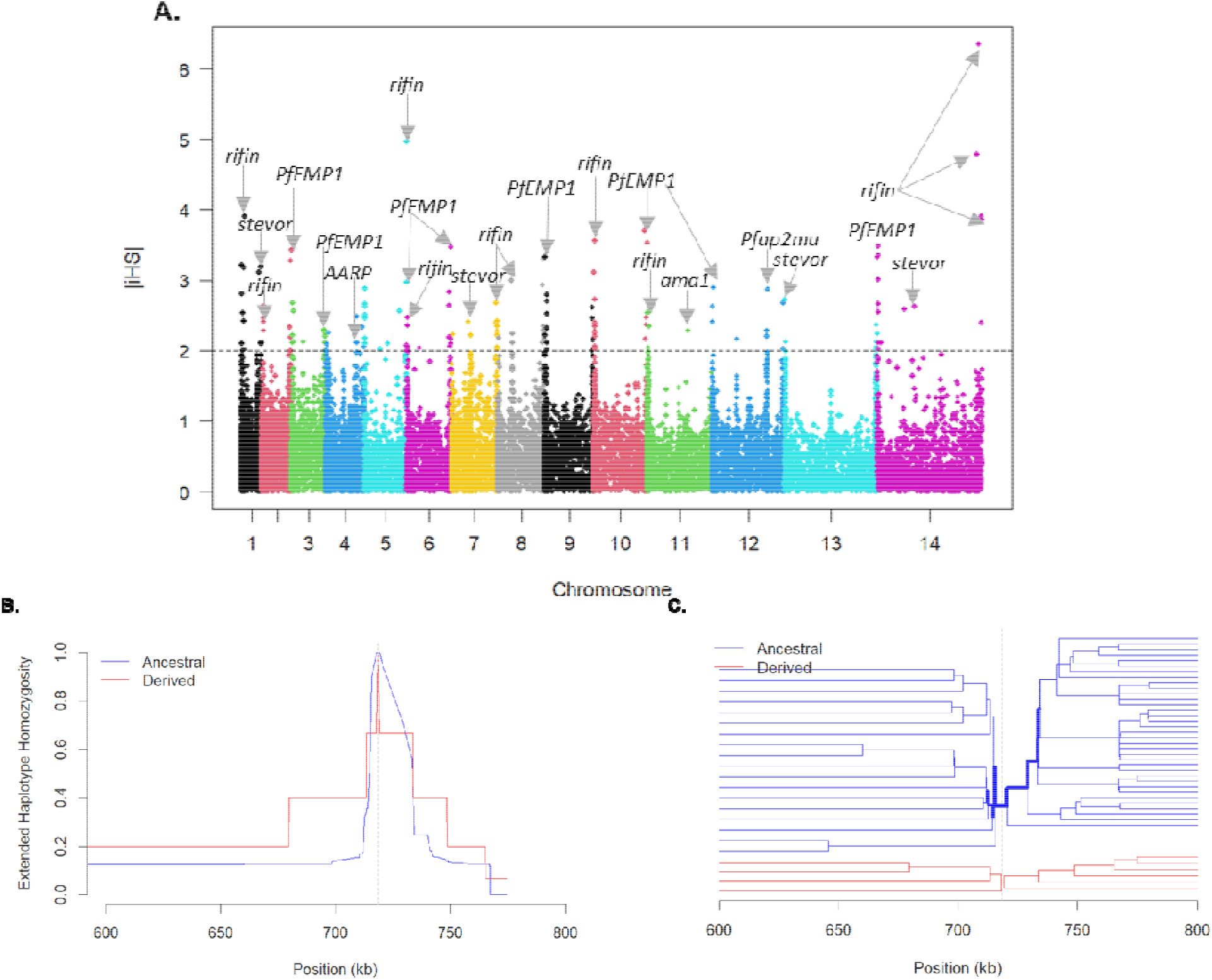
Genome-wide scans for selection signals A) Integrated Haplotype Score (iHS) Manhattan plot with individual chromosomes identified by alternate coloring of their SNPs. Genes with high scoring SNPs (iHS>2) suggest recent positive directional selection. **B)** Extended haplotype homozygosity (EHH) plot showing the homozygosity of the most frequent extended haplotype around the SNP 160**N**. X-axis of the EHH plot shows the upstream and downstream genomic coordinates from the locus of interest. Homozygosity scale is shown on the y-axis, ranging from 0 to 1 (0 implying no homozygosity and 1 complete homozygosity). Haplotype bifurcation diagrams in panel (**C**) showing breakdown of linkage disequilibrium in individuals carrying the ancestral and derived allele of the polymorphism 160**N**. This is bidirectional with the root representing a core SNP (at 160**N**) depicted by a vertical dotted line. Thickness of the line corresponds to the number of samples with shared haplotype.

## Discussion

*P. falciparum* is continuously evolving to become resistant to antimalarials. There is a particular concern for this selection to be accelerated in low transmission regions, where decreased immunity may lead to increased symptomatic cases which eventually increase drug treatment and pressure on the parasite population.^25,26^ In addition, the high number of monogenomic infections and clonal recombination of highly related parasites in low transmission settings decreases competition with wildtype strains, compared to high transmission settings where within host competition, high polyclonality, and genetic diversity with significant recombination among diverse clones is thought to delay the spread of drug resistance.^38,39^ Choma District of Zambia is such a low transmission region yet little longitudinal data exist on molecular markers of antimalarial drug resistance or genomic signatures of how the parasite population may be changing in the face of control efforts. Here, we provide evidence through longitudinal molecular surveillance, supported by genomic analysis of selection of temporal changes in antimalarial resistance that are concerning for both antimalarial therapies with ACTs and chemoprevention with antifolate drugs. As endemic countries like Zambia successfully approach pre-elimination, implementation of genomic surveillance can clarify how current control methods affect parasite populations and monitor for the emergence and spread of antimalarial drug resistance.^40^

Zambia was the first African country to adopt the use of ACTs with the use of artemether-lumefantrine (AL) in 2003, highlighting the importance of monitoring ART-R mutations that arise *de novo* in addition potential importation of resistant mutations,^41^ and resistance mutations to the partner drugs leading to treatment failure. The country is landlocked and shares a border with countries where malaria transmission is high, which could contribute to the potential for parasite importation.^41^ Another concern is that the existing practice of the use of different antimalarial drugs for mass drug administration (MDA)^42^ could lead to the natural selection of drug-resistant *P. falciparum* strains.^43^ Genotyping drug resistance markers from longitudinal samples collected from decreasing parasite population from low transmission setting provides the opportunity to assess how past and current intensified malaria control have affected the emergence and spread of resistant strains, providing an early warning system for drug resistance.

None of isolates in this study carried WHO validated or candidate mutations R561**H**, A675**V** and C469**Y** found commonly in east Africa.^18,44^ However, one sample had a WHO validated artemisinin partial resistance marker Kelch 13 R622**I**. Identification of a Kelch 13 R622**I** mutation in this low transmission setting could mean one of two things: 1) parasites have emerged in the region bearing ART-R mutations; or 2) parasites have been imported that have this mutation. This study cannot answer that question, but supports the need for intensified surveillance. A recent study from Ethiopia^5,30^ and Eritrea^7^ have also shown that the Kelch 13 622**I** mutation can co-exist with diagnostic-resistant histidine-rich proteins 2 and 3 (*Pfhrp2/3*) deleted parasite strains in low transmission settings. Moreover, we found five novel *de novo* mutations C532**S**, A578**S**, Q613**E**, D680**N** and G718**S** within the Kelch 13 propeller domain suggesting that parasites are under pressure due to continued use of ACT. This warrants close monitoring of the emergence and spread of mutations associated with ART-R.

Also concerning for artemisinin efficacy in Zambia are the findings of increasing AP2MU mutations. The 160**N** mutation was associated with artemisinin resistance in Africa.^33,34^ Our data suggests both an increasing prevalence in the community (**Fig 1**) and genomic signatures of positive selection at this site are consistent with selection due to drug pressure (**Fig 6**). This supports the finding of directional selection at this locus in the African *P. falciparum* population.^45^ This combination suggests that current ACT practices are impacting the parasite population but further investigation into the phenotypic association between this mutation and antimalarial resistance is needed. Other posited markers of ART-R, such as UBP1 and ATP6, did not have concerning signatures in our analysis.

Our genomic analysis of *P. falciparum* parasites from southern Zambia identified known resistance mutations in DHFR, DHPS, MDR1, and other genes. This was not surprising since mutations in these genes have been previously documented within Zambia.^46^ While AL retains its efficacy against uncomplicated *P. falciparum* malaria in Zambia, the upsurge of MDR1 N**F**D haplotype in recent years associated with lumefantrine resistance calls for close monitoring of mutations associated to partner drugs.^47^ We did not find any evidence for mutations that would affect amodiaquine efficacy. Given the alternating selective pressure between lumefantrine and amodiaquine in neighbouring countries like Tanzania, the persistence of amodiaquine sensitivity is important for Zambia’s malaria control program.

The effectiveness of antimalarials used for chemoprevention is also compromised by antimalarial resistance. SP is a widely used drug for IPTp and IPTi in southern Zambia.^23^ Persistence and potential increases in the IRNGE genotypes threaten the utility of this drug for malaria prevention. Our findings support other studies in Africa that have also found that the prevalence of SP resistance mutations to be high.^13,48^ However, none of the parasites in the current study carried DHFR 164**L** and only 2 samples carried the DHPS 581**G** mutation that confers high-level resistance to sulfadoxine. The WHO-recommended thresholds for the withdrawal of SP in IPTp are when DHPS 540**E** > 95% and 581**G** > 10%, which this area of Zambia has not yet reached.^49^

Interestingly, WGS showed evidence of recent positive selection, among variable antigen genes often involved in immune evasion. We previously showed that there is a high degree of clonal transmission in this area through evaluation of identity by descent (IBD).^50^ Reductions in effective population size and increasing clonal transmission may also impact these hypervariable genes identified under selection in the iHS analysis (**Fig 6A**). These signatures of selection may be indicative of continued strong control in Southern Zambia.

Our study provides insights into the prevalence of drug resistant genetic markers and how current and past control efforts have affected malaria parasite populations through longitudinal molecular surveillance and genomic analysis for signatures of selection. The presence of mutations associated with reduced efficacy for malaria therapy with ACTs and malaria chemoprevention with SP are concerning for malaria control efforts in the region. Coordinated efforts for widespread longitudinal molecular surveillance, combined with appropriate sampling design and collection of travel histories, will be essential for achieving malaria elimination. This study provides additional data for developing a starting point to help engage stakeholders and provide preliminary data and hypotheses for ramping up malaria molecular surveillance in low transmission settings like southern Zambia. Therefore, continued monitoring of temporal changes in frequency and distribution of antimalarial drug resistance across Zambia is warranted.

## Methods

### Sample collection

Samples that underwent MIP genotyping were dried blood spots (DBS, n=455) collected between 2012 and 2018 from symptomatic RDT-positive cases at 8 health centres within the catchment area of Macha Hospital in Choma District, Southern Province that encompasses 2,000 km^2^. In addition, 28 samples from persons presenting with uncomplicated malaria in 2019 to Mapanza Rural Health Center within the catchment areas of Macha Hospital underwent WGS.

This work was approved as part of the Southern and Central Africa International Center of Excellence for Malaria Research (ICEMR) by the Tropical Diseases Research Centre, Ndola Ethics Review Committee (Ref No: TDRC/ERC/2010/14/11) and the Johns Hopkins Bloomberg School of Public Health Institutional Review Board (IRB # 3467). Sequence analysis using parasite genomes from de-identified samples and data were deemed nonhuman subjects of research at the University of North Carolina at Chapel Hill (NC, USA) and Brown University (RI, USA).

### MIP sequencing of drug resistance loci

DNA was extracted from each of the 455 DBS specimens with a Chelex-Tween protocol.^51^ Parasitemia was assessed using quantitative PCR with probes targeting the *P. falciparum ldh* sequence.^52^ All samples were then genotyped using a MIP panel (n=815) targeting known drug resistant SNP mutations and select coding sequences in 14 genes across the *P. falciparum* genome.^53^ MIP capture and library preparation were done as previously described.^54^ Sequencing was conducted using an Illumina NextSeq 550 instrument (150 bp paired-end reads) at Brown University (RI, USA).

### MIP data analysis and estimating drug resistance prevalence

Processing of sequencing data and variant calling was done using MIPtools (v0.19.12.13; https://github.com/bailey-lab/MIPTools), a suite of computational tools designed to process MIP Illumina sequencing and provide haplotype and variant calls. Briefly, after demultiplexing samples, raw reads from each captured MIP, identifiable using unique molecular identifiers (UMIs), were used to reconstruct sequences using MIPWrangler, and variant calling was performed on these samples using freebayes.^55^ Biallelic, variant SNP positions were retained for analysis. Variants were annotated using the 3D7 v3 reference genome. To reduce false positives due to PCR and sequencing errors, the alternative allele (SNP) must have been supported by more than one UMI within a sample with minimum coverage of 10x, and the alternative allele must have been represented by at least 10 UMIs across the entire population.

The prevalence of point mutations for each drug resistance markers was calculated as p=x/n*100, where p = prevalence, x = number of infections with mutant alleles, n = number of successfully genotyped infections. Mutant combinations were plotted and visualized using *UpSet* Package in R.^56^ Where appropriate, all outputs were visualized using the ggplot2 package in R.

### Whole genome sequencing and tests for signatures of selection

The 28 *P. falciparum* DBS samples were punched and extracted using a QIAcube HT instrument and QIAmp 96 DNA kit (Qiagen: Hilden, Germany) using an optimized high throughput genomic DNA (gDNA) extraction protocol.^57^ gDNA quantity and quality were assessed using Qubit 1x dsDNA High Sensitivity Assay (ThermoFisher Scientific: Waltham, MA) and ScreenTape on the Agilent TapeStation 4150 (Agilent Technologies: Santa Clara, CA), respectively. A 4-plex hybrid capture method using SeqCap EZ custom probes^58^ was then used to selectively enrich *P. falciparum* DNA from DBS samples containing host DNA due to the low parasitaemia in these samples. All WGS samples were sequenced using NovaSeq 6000 technology system (Illumina: San Diego, CA), and paired-end reads were mapped to the *P. falciparum* 3D7 reference genome using BWA-MEM 0.7.17. Variants and down streaming filtering were called using GATK v4.1.4.1^21^ following best practices (https://software.broadinstitute.org/gatk/best-practices). From the unphased VCF file, we calculated selection using the standardized integrated haplotype score (iHS) and estimated integrated extended-haplotype homozygosity (EHH) values between alleles at a given SNP. All associated iHS and EHH calculations were carried out using the R-package rehh (version 2.0.4). (https://cran.r-project.org/web/packages/rehh/index.html).

## Data Availability

All sequencing data available under Accession no. SAMN29983042 - SAMN29983315 at the Sequence Read Archive (SRA) (http://www.ncbi.nlm.nih.gov/sra), and the associated BioProject alias is PRJNA862735.

## Funding

This work was supported by the National Institute for Allergy and Infectious Diseases (U19AI089680 to WJM, K24AI134990 to JJJ, R01AI139520 to JAB).

## Author contributions

AAF, JJJ and JAB conceived the study. CH, TK, TS and HH were responsible for DNA extraction and molecular diagnosis of *Plasmodium falciparum* infections. AAF did MIP initial data analyses. AAF, JJJ and JAB wrote the manuscript and undertook final data analyses and interpretations. All authors contributed to the writing of the manuscript and approved the final version.

## Acknowledgments

We thank the participants of the study.

## Supplementary Materials

### Supplementary Tables

*Supplementary tables are compiled into a single file for ease of viewing*

**Table S1. Prevalence of key antimalarial resistance mutations by site and year**.

n = number of samples successfully sequenced per loci, NA = Not applicable, mutant = both pure and mixed mutants.

See uploaded file.

**Table S2. Loci under selection as detected by iHS**. A total of 164 loci under selection identified across 14 chromosomes of *P. falciparum* genome. We define a genomic region as being under selection if it contains at least two extreme markers with the |iHS|□>□2 (top 1% of the expected distribution).

See uploaded file.

### Supplementary figures

**Figure S1.**
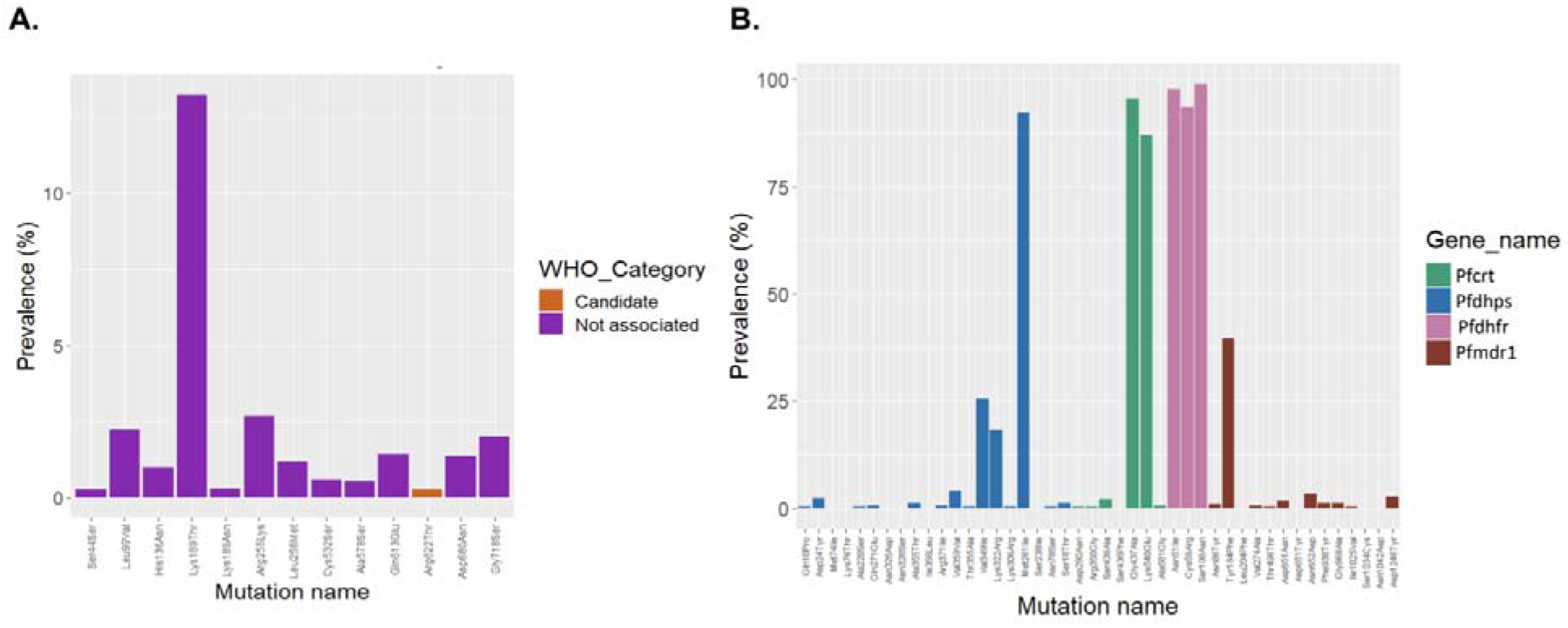
Antimalarial Resistance Polymorphisms. Panel A shows the prevalence of K13 mutations in the genotyped samples. Candidate artemisinin partial resistance mutations are shown in orange, while other mutations are shown in purple. Panel B shows the prevalence of other antimalarial resistance profiles. Mutations in key genes are colored differently.

**Figure S2.**
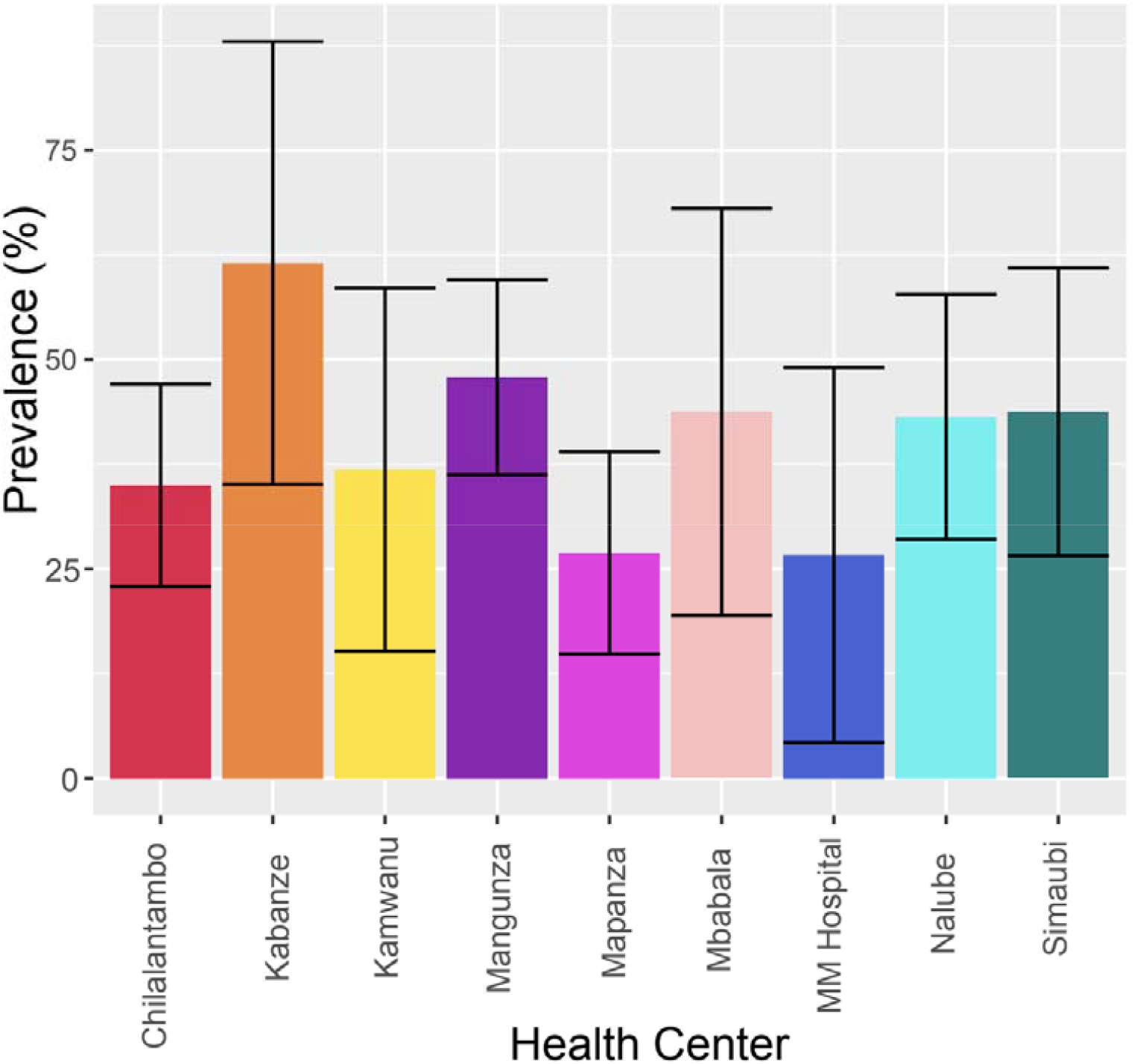
Spatial trends of MDR1 N86/184F/D1246 haplotype in Choma district, Southern Zambia. Bar plot showing the mean values with standard deviation as error. Error bars on both sides. Colors indicate health facilities.

## Notes

### Competing Interest Statement

The authors have declared no competing interest.

### Author Declarations

This work was approved as part of the Southern and Central Africa International Center of Excellence for Malaria Research (ICEMR) by the Tropical Diseases Research Centre, Ndola Ethics Review Committee (Ref No: TDRC/ERC/2010/14/11) and the Johns Hopkins Bloomberg School of Public Health Institutional Review Board (IRB # 3467).

